# Predicting SARS-CoV-2 variant spread in a completely seropositive population using semi-quantitative antibody measurements in blood donors

**DOI:** 10.1101/2022.06.16.22276483

**Authors:** Lewis F Buss, Carlos A Prete, Charles Whittaker, Tassila Salomon, Marcio K. Oikawa, Rafael H. M. Pereira, Isabel C. G. Moura, Lucas Delerino, Rafael F. O. Franca, Fabio Miyajima, Alfredo Mendrone-Junior, César de Almeida Neto, Nanci A. Salles, Suzete C. Ferreira, Karine A. Fladzinski, Luana M. de Souza, Luciane K. Schier, Patricia M. Inoue, Lilyane A. Xabregas, Myuki A. E. Crispim, Nelson Fraiji, Luciana M. B. Carlos, Veridiana Pessoa, Maisa A. Ribeiro, Rosenvaldo E. de Souza, Anna F. Cavalcante, Maria I. B. Valença, Maria V. da Silva, Esther Lopes, Luiz A. Filho, Sheila O. G. Mateos, Gabrielle T. Nunes, David Schlesinger, Sônia Mara Nunes da Silva, Alexander L. Silva-Junior, Marcia C Castro, Vítor H. Nascimento, Christopher Dye, Michael P Busch, Nuno R Faria, Ester C Sabino

## Abstract

**Background:** SARS-CoV-2 serologic surveys estimate the proportion of the population with antibodies against historical variants which nears 100% in many settings. New analytic approaches are required to exploit the full information in serosurvey data.

**Method:** Using a SARS-CoV-2 anti-Spike (S) protein chemiluminescent microparticle assay, we attained a semi-quantitative measurement of population IgG titres in serial cross-sectional monthly samples of routine blood donations across seven Brazilian state capitals (March 2021-November 2021). In an ecological analysis (unit of analysis: age-city-calendar month) we assessed the relative contributions of prior attack rate and vaccination to antibody titre in blood donors. We compared blood donor anti-S titre across the seven cities during the growth phase of the Delta variant of concern (VOC) and use this to predict the resulting age-standardized incidence of severe COVID-19 cases.

**Results:** On average we tested 780 samples per month in each location. Seroprevalence rose to >95% across all seven capitals by November 2021. Driven proximally by vaccination, mean antibody titre increased 16-fold over the study. The extent of prior natural infection shaped this process, with the greatest increases in antibody titres occurring in cities with the highest prior attack rates. Mean anti-S IgG was a strong predictor (adjusted R2 =0.89) of the number of severe cases caused by the Delta VOC in the seven cities.

**Conclusions:** Semi-quantitative anti-S antibody titres are informative about prior exposure and vaccination coverage and can inform on the potential impact of future SARS-CoV-2 variants.

**Summary:** In the face of near 100% SARS-CoV-2 seroprevalence, we show that average semi-quantitative anti-S titre predicted the extent of the Delta variant’s spread in Brazil. This is a valuable metric for future seroprevalence studies.

## Introduction

Serologic surveys estimate the proportion of a population with detectable antibodies against SARS-CoV-2 to infer cumulative incidence of infection or vaccination coverage (e.g. [1]). This approach was useful early in the pandemic to estimate approximate total infections, important for epidemic modelling and determining fatality ratios. However, as population exposure to SARS-CoV-2 antigens, through infection or vaccination, reaches 100%, so too will seroprevalence (e.g. [2]), making this indicator meaningless. The interpretation is further limited by successive emergence of SARS-CoV-2 variants of concern (VOC) [3], with increasing transmissibility [4,5] and partial immune escape [6,7], that continue to cause epidemics in populations with high documented immunologic exposure [8,9].

The semi-quantitative output, reported as the ratio of signal-to-cut off (S/C), arbitrary units (AU)/mL, or binding antibody units (BAU)/ml, of serological assays reflects IgG or total antibody titres and thus provides additional information over a binary positive *versus* negative interpretation. The average population S/C value will be some function of the extent and timing of prior natural infection, as well as vaccine coverage, type and timing. Together, these factors will shape population patterns of immunity towards SARS-CoV-2 infection and severe COVID-19 disease [10]. Previous work has shown neutralising antibody levels to be highly predictive of protection from SARS-CoV-2 symptomatic infection[11]. As such, higher average semi-quantitative anti-S levels might predict a lower incidence of severe cases caused by a new variant introduced into a seropositive population.

Here we test this hypothesis using serial cross-sectional semi-quantitative anti-Spike (S) protein measurements from across Brazil in 2021 when the Gamma [6], Delta [12] and Omicron [13] VOCs successively replaced one another. We assess the extent to which vaccination and prior infection contributed to measured population anti-S IgG levels, and the degree to which these variables predicted the incidence of severe COVID-19 cases caused by the Delta VOC.

## Methods

### Study design and blood donor sampling strategy

The blood donor sampling strategy has been reported in detail previously [9,14]. Briefly, we aimed to test 850 blood donation samples each month from public blood banks in the seven participating Brazilian cities (São Paulo, Rio de Janeiro, Manaus, Recife, Fortaleza, Curitiba, Belo Horizonte). Starting from the second week of each month, we selected consecutive samples among all donations (in Manaus) or within city neighbourhoods to achieve spatial representativity (remaining cities). Sample selection proceeded until quotas were filled or the available samples were exhausted. Sampling spanned donations from April 2021 to November 2021 in all seven cities. In addition, we tested samples from November 2020 in Manaus (previously reported in [14]) and March 2021 in Recife.

### SARS-CoV-2 serology assay

We used a chemiluminescent microparticle immunoassay (CIMA, AdviseDx, Abbott) that detects IgG antibodies against the SARS-CoV-2 spike (S) protein. At a threshold of 50 S/C units, we have previously shown this assay to have a sensitivity of 94.0% in 208 non-hospitalized PCR-positive convalescent individuals, and its specificity is >99% [15].

### Secondary data sources

The numbers of vaccine doses administered were taken from the OpenDataSUS [16]. Population denominators were projected estimates for 2021 based on the 2010 Brazilian census [17].

We analysed three different sources for COVID-19 cases. First, we retrieved all severe acute respiratory syndrome cases (SARI, confirmed COVID-19 and unknown aetiology) and deaths from the SIVEP-Gripe [16]. Total SARI is less affected by underreporting [18,19], but only reflects severe cases. Second, we retrieved the total number of confirmed COVID-19 cases (irrespective of severity) reported by the Brazilian Ministry of Health. The official MoH case counts should be treated with caution as they are heavily influenced by access to testing and are published by date of reporting not date of symptom onset[18]. Finally, we further obtained the time series of SARS-CoV-2 test positivity in a network of pharmacies in São Paulo city (https://mendelics.com.br/).

To determine the relative abundance of SARS-CoV-2 variants we retrieved metadata from all SARS-CoV-2 genomes deposited on GISAID [20] between March 2020 and March 2022 in the seven states. We grouped SARS-CoV-2 lineages into Omicron, Delta, Gamma, and wildtype and summarized as weekly lineage counts per location.

### Data analysis and statistics

We fit a multinomial model to weekly variant counts with calendar time as the predictor variable and a two-knot cubic spline [4] using the nnet package [21] in R (version 4.1.2). Using the model, we arbitrarily defined periods of variant dominance in each state as beginning from the week when each variant first reached 10% relative abundance.

We calculated the incidence of SARI cases and deaths using the projected population size for the seven cities. For total cases from the MoH, there was an artefactual increase in the number of reported cases in a small number of weeks, reflecting changes in reporting, not transmission. To remove this effect but preserve the overall shape of the epidemic curve, we excluded four weeks for Curitiba and one for Recife where the case counts were >10 times the median for those cities.

We first calculated monthly IgG anti-S seroprevalence estimates with exact binomial 95% confidence intervals (95%CI) for all cities using the manufacture’s threshold of 50 S/C units to define a positive reading. We then calculated the geometric mean of the semi-quantitative anti-S IgG S/C readings from all blood donations for each month and location. S/C scales vary between serology assays, and there is no direct biological interpretation of these values. As such, we followed the approach of Khoury et al and Earle et al [11,22], and standardized the mean anti-S IgG S/C values against a convalescent cohort. We calculated the ratio of the mean monthly S/C readings to the mean S/C value seen in a cohort of 245 convalescent samples collected following wild type infection in 2020. This cohort has been described in detail previously [9,14,23].

In primary studies of vaccine efficacy, the convalescent-normalized mean anti-S S/C induced post-vaccination correlate strongly with protection against PCR-confirmed symptomatic infection with wild type SARS-CoV-2 [11,22]. We fit a linear regression to results from seven primary vaccine studies (data from [22]), and used this to estimate convalescent-normalized mean anti-S S/C that equate to 65%, 75% and 85% vaccine efficacy.

We next explored the association between convalescent-normalized anti-S S/C, vaccination, and prior attack rate. We exploited the variation in vaccination timing across the blood donor age range (15-69yr), where older individuals were vaccinated before younger individuals. We built a multivariate linear regression of monthly convalescent-normalized mean S/C in 10-year age groups on vaccine coverage with the first dose, second dose and prior attack rate by Dec 2020 (as estimated previously [14]) in each city. We built a null model containing all three variables and no interaction terms. We then added interaction terms between attack rate and first dose coverage, and attack rate and second dose coverage. We selected the best performing model using the Bayesian Information Criterion.

Finally, we assessed the predictive value of convalescent-normalized mean anti-S S/C to predict the spread of the Delta VOC across the seven cities. For a two-month period starting from the day Delta reached 10% dominance in each city, we calculated age-standardized SARI incidence in the blood donor age range. Standardization was with the direct method taking the population of São Paulo as the reference. We then regressed age-standardized SARI incidence on vaccine coverage (first and second dose separately) and convalescent-normalized mean anti-S S/C.

### Data availability

Continuing our previous serological data sharing initiative [8] all raw serological data are available at https://github.com/CADDE-CENTRE.

### Ethics

This project was approved by the Brazilian national research ethics committee, CONEP CAAE -30178220.3.1001.0068.

## Results

### SARS-CoV-2 infection, vaccination, and seroconversion across seven Brazilian capitals

The seven capital cities (Belo Horizonte, Curitiba, Fortaleza, Manaus, Recife, Rio de Janeiro, and São Paulo) are located across four of Brazil’s five macro-regions (Figure 1A). Vaccination was rolled out across the seven cities at the beginning of 2021. Among residents in the blood donation-eligible age range (15-69yr), coverage with the first dose reached >75% across all cities by the end of the study period. Coverage with the second dose was also high by this point (Figure 1B). The share of vaccine types is shown in Figure 1C. Importantly, Sinovac accounted for ∼25% across all cities, and induces both anti-S and anti-N antibodies. For this reason, the presence of anti-N antibody cannot be used to distinguish natural infection from vaccine-induced seroconversion in the Brazilian context.

**Figure 1.**
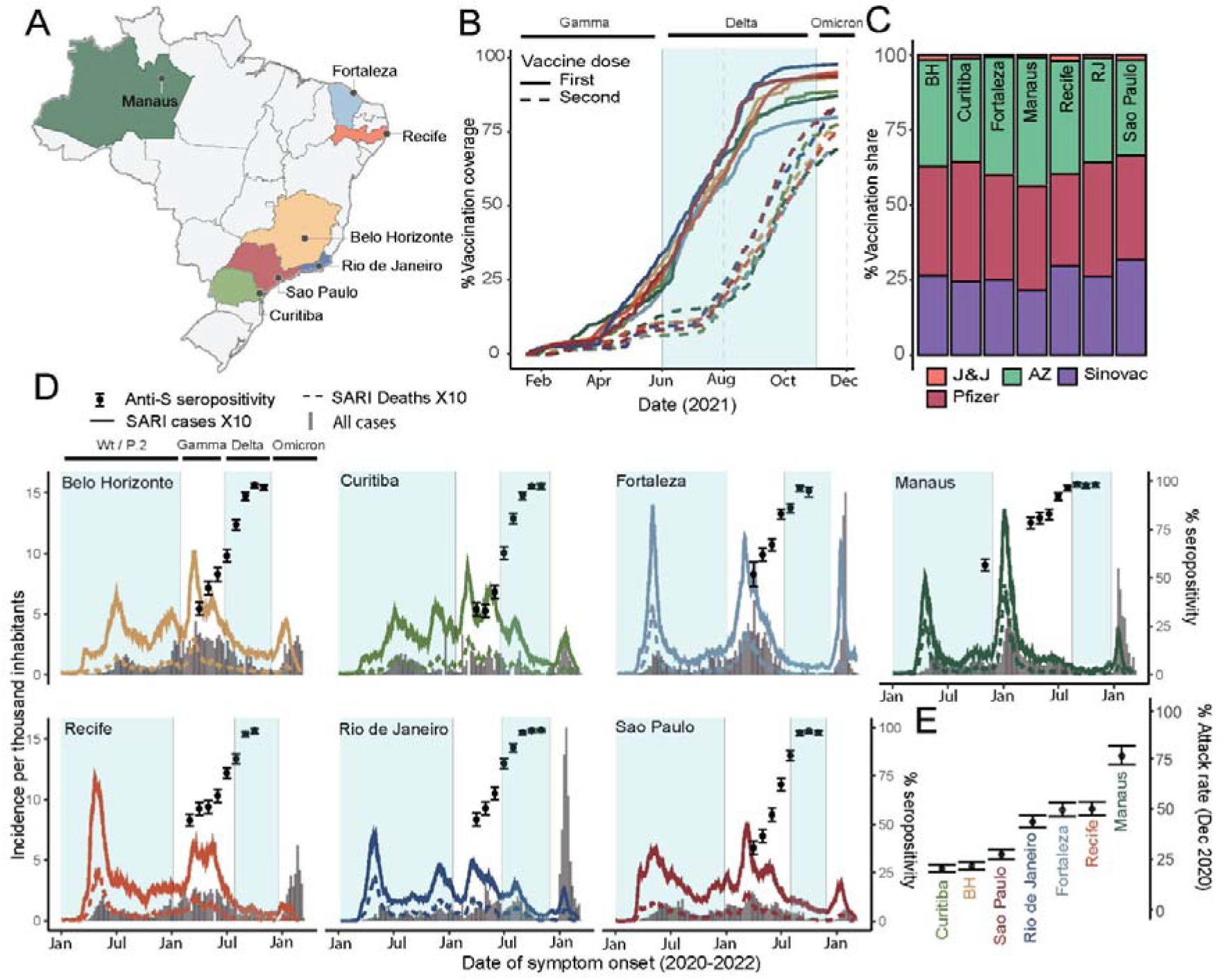
SARS-CoV-2 infection and vaccination across seven Brazilian capital cities. A – location of the seven state capitals with participating blood donation centres. B – proportion of the population within the donation-eligible age group (15-65yr) having received one or two doses of SARS-CoV-2 vaccine in each of the seven cities. C – the relative share of vaccine types cumulative through Dec 2021. BH – Belo Horizonte, RJ – Rio de Janeiro. J&J – Johnson and Johnson, AZ – AstraZeneca. Vaccination data was extracted from OpenDataSUS (https://opendatasus.saude.gov.br/). D – incidence of cases and deaths due to severe acute respiratory syndrome (SARI, data are from SIVEP-Gripe national reporting system) multiplied by 10 (for visualization alongside total cases) and total cases reported by the Brazilian Ministry of Health (https://covid.saude.gov.br/). Periods of variant dominance start from the date where each variant reached a 10% share of all sequences deposited on GISAID (https://www.gisaid.org/) based on predictions from a multinomial model fit to this data (Figures S1). Anti-S seropositivity is calculated based on monthly blood donor samples and are shown with exact 95% binomial confidence intervals (error bars). E – Estimated cumulative attack rate in Dec 2020 based on anti-N serosurveillance in the same blood donor population (Prete et al. Research Square, 2022). 95% confidence intervals are shown as error bars.

Distinct SARS-CoV-2 epidemics, in both shape (Figure 1D) and size (Figure 1E), have occurred across these cities. For example, the cumulative attack rate (inferred from seroprevalence) in Dec 2020, prior to the Gamma-driven second wave and prior to the rollout of vaccination campaigns in Brazil, ranged from 20.3% (95% confidence interval, 95% CI, 18.6% to 22.3%), in Curitiba, to 76.3% (95% CI 72.1% to 81.4%) in Manaus (Figure 1E) [14].

Whilst all cities experienced a large Gamma-dominated peak in cases and deaths (Figure 1D, Supplemental Figure S1), the subsequent period of Delta dominance was not marked by similarly significant epidemics. Indeed, incidence following Delta’s introduction was characterised by persistently low (Manaus, Fortaleza, Recife), falling (Belo Horizonte, São Paulo) or falling with a small initial increase (Rio de Janeiro, Curitiba) incidence of cases and deaths (Figure 1D). There was a variable peak in the number of reported cases with the introduction of Omicron, but a negligible increase in deaths: only 3.7% of deaths were reported in this period.

We tested on average 780 (range 247-997) blood donation samples (per month per location) for anti-S IgG. The proportion of donors testing positive on this assay (S/C > 50) increased steadily over 2021, reaching >95% in all cities (95.1% in Fortaleza to 98.6% in Recife) by November 2021.

### Convalescent-normalized mean S/C in blood donors, vaccination coverage and prior attack rate

The monthly convalescent-normalized mean anti-S readings are presented in Figure 2A (raw data in Figure S2 and S3). S/C values corresponding to 65%, 75% and 85% hypothetical vaccine efficacy against wild type are shown as dashed lines (Figure 2). Due to several limitations (see Discussion) these should not be interpreted as the true population protection in the cities at this these time points.

**Figure 2.**
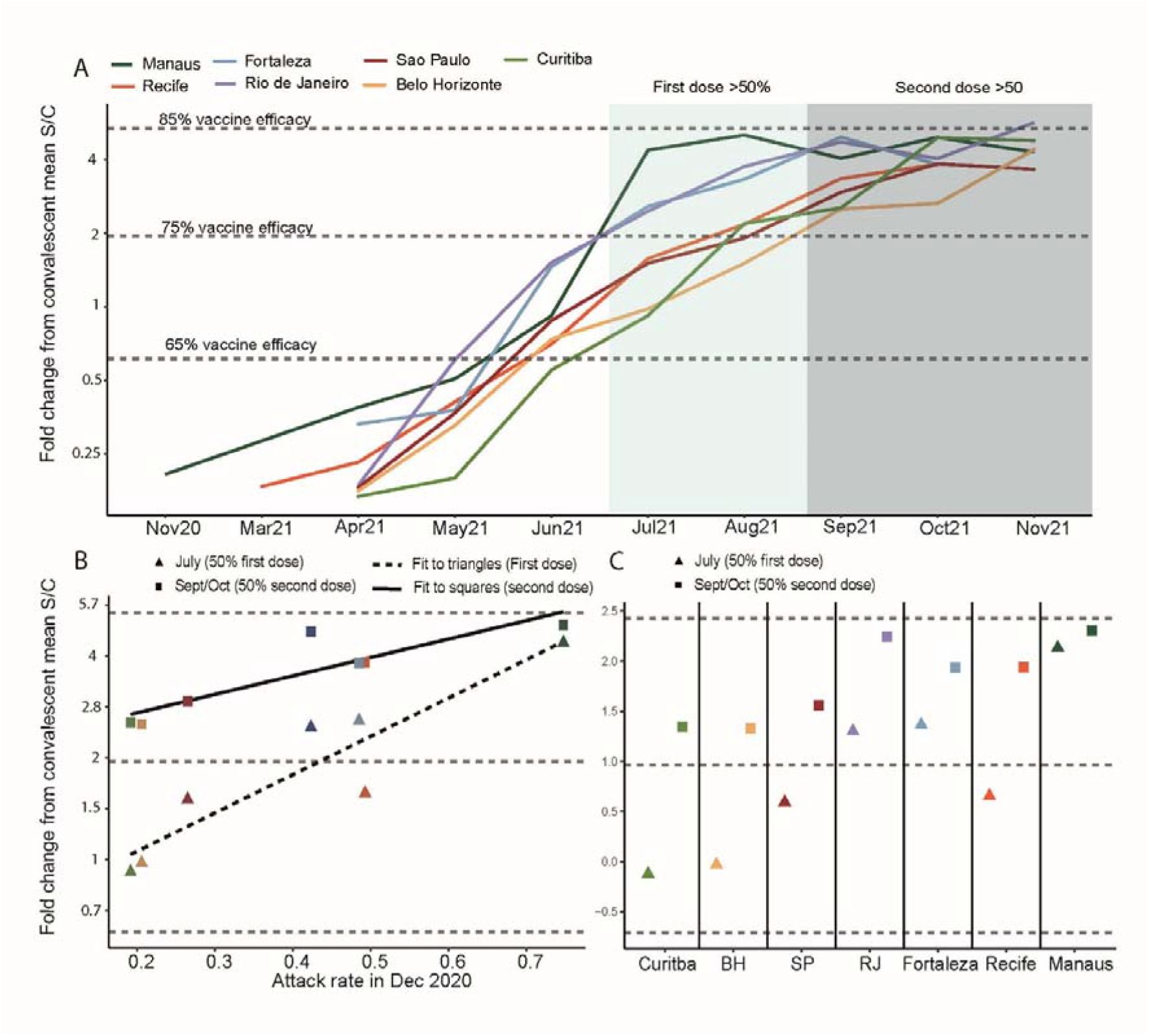
Monthly convalescent-normalized mean antibody titre in blood donors across seven Brazilian cities. A - Mean anti-S IgG antibody titre in blood donors normalized against mean convalescent anti-S IgG level shortly following infection. B – Convalescent-normalized anti-S IgG when first dose coverage reached 50% in all cities (July 2021, triangles) and second dose vaccination reached 50% (September or October, squares) against estimated attack rate in December 2020 (Prete et al. Research Square, 2022). C – symbols as in B but grouped by city on x-axis. Vaccine efficacy estimates (horizontal dashed lines) are based on the relationship between convalescent-normalized mean anti-S IgG following vaccine administration and protection against PCR-confirmed symptomatic infections (as described in Earle et al. 2021).

Mean antibody concentration increased 16-fold between May and August 2021 (Figure 2A). During this period, the first vaccine doses were administered in the blood donor age range (Figure 1B). This appears to have been the proximal factor driving the dramatic increase in antibody S/C values. Coincident with this there was also a variable amount of natural infection occurring across the seven cities (Figure 1D), and the contribution of the two cannot be completely separated.

Prior attack rate influence the S/C reached when vaccine coverage was at 50% (Figure 2B and 2C). In Manaus, where the prior attack rate was highest, administration of the first dose was associated with a large increase in antibody levels (Figure 2B), whereas the incremental change following the second dose was small (Figure 2C). In the other cities, the second dose was associated with larger increases in antibody titres. Consistent with these observations, the best fitting model for convalescent-normalized antibody level included interactions between prior attack rate and first and second dose coverage (Table 1). Assuming a prior attack rate of zero, each 10% increase in first dose vaccination resulted in a 1.17-fold (95% CI 1.09 to 1.26) increase in convalescent-normalized mean antibody levels. The magnitude of this increase was 1.03 (1.01-1.04) greater for each 10% increase in the prior attack rate. In contrast, the effect of the second dose coverage was 0.97 (0.95-0.99) smaller for each 10% increase in the prior attack rate.

**Table 1.** Best performing linear regression model of convalescent-normalized monthly antibody anti-S titres.

| **Model terms** | **Point estimate (95% CI)** | **p-value** |
| --- | --- | --- |
| Per capita x 10 * | Fold change from mean<br>convalescent |  |
| First dose coverage | 1.18 (1.10-1.26) | <0.001 |
| Second dose coverage | 1.15 (1.07-1.24) | <0.001 |
| Attack rate | 1.07 (0.99-1.16) | 0.10 |
| First dose x attack rate | 1.03 (1.01-1.05) | 0.002 |
| Second dose x attack rate | 0.97 (0.95-0.99) | <0.001 |

An increase of 1 unit corresponds to a 10% increase in coverage or attack rate

### Anti-S IgG levels in blood donors as predictor of Delta’s epidemic penetrance

The Delta VOC first reached a 10% share of genomes on GISAID in Curitiba (19/06/2021) and last in Manaus (16/08/2021). There was a strong relationship between age-standardized SARI and anti-S S/C (Figure 3), with 89% of the variance in epidemic size explained. A weaker relationship existed between population coverage with the first and second vaccine doses (Figure 3), with 49% and 39% of the variance explained by these variables, respectively.

**Figure 3.**
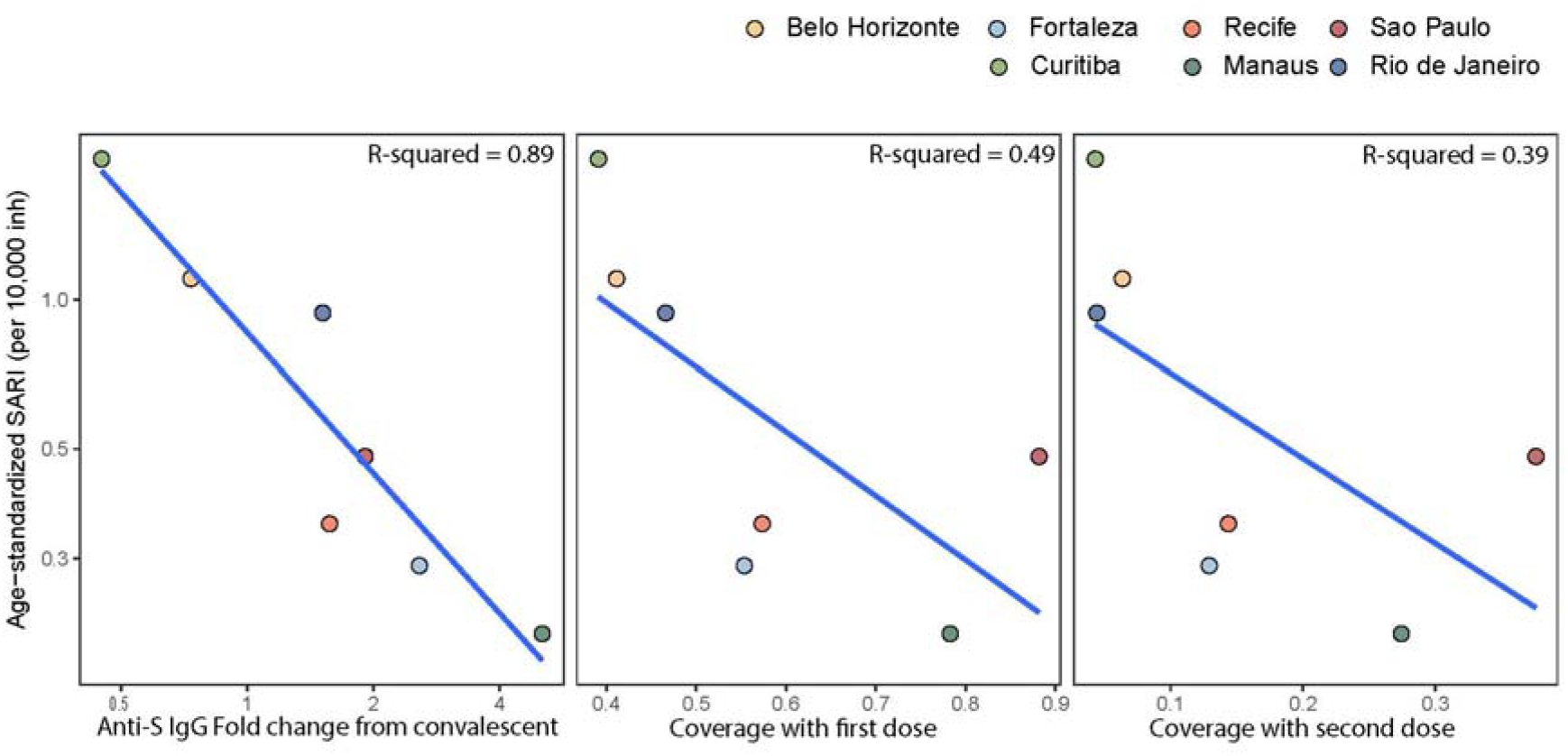
Predictors of epidemic size of the Delta growth phase across seven Brazilian cities. Convalescent-normalized mean anti-S IgG signal-to-cutoff was calculated at the month when Delta reached 10% dominance in each of the cities (range 19/06/2021 in Curitiba to 16/08/2021 in Manaus). Percentage coverage with the first and second doses was also calculated up to (and inclusive of) the month of 10% dominance in each city. Total severe acute respiratory syndrome (SARI) cases, within the age range of blood donors (15-65) were age standardized using the direct method and the age structure of São Paulo as the reference population. A two-month period starting from the date of 10% dominance was used to calculate epidemic size. R-squared terms are from separate simple linear models fit to the seven points shown on the figure.

## Discussion

We present serial cross-sectional anti-S IgG results in blood donors across seven Brazilian state capitals in 2021. Nearly all donors were seropositive by the end of the year. While vaccination was a key factor that increased quantitative antibody levels, historical attack rates determined the dynamics and scale of these increases. Specifically, first dose coverage was associated with a steeper increase in antibody S/C in cities with higher prior attack rates. By contrast, the second dose was associated with a smaller increment in antibody when prior attack rate was higher. This is consistent with individual-level data [24] showing that the first vaccination dose induces high anti-S S/Cs in individuals previously infected by SARS-CoV-2, and that these levels were similar to double-dosed immune naïve people. By contrast, the second dose produced a minimal increment in antibody level among convalescents but had a larger effect on those not previously infected. Our results show that semi-quantitative anti-S levels in blood donors varied as a function of the complex combination of immunizing events within these seven Brazilian urban populations.

An important question is whether average population anti-S S/C readings have an epidemiologic interpretation – specifically, whether they can they be used to predict morbidity from emerging variants. This might be expected to be the case, given the strong relationship between mean anti-S binding antibody level induced by vaccination and the group-level protection against wild type infection and COVID-19 disease penetrance [22]. Furthermore, anti-S level correlate strongly with neutralizing antibody titres [23] and these in turn are predictive of group-level vaccine efficacy [11,22]. Indeed, our results showed that, during the growth phase of the Delta VOC in Brazil, the incidence of SARI cases was strongly correlated with antibody level measured in blood donors.

This finding is further supported by individual level data [25]. The risk of infection with the Delta VOC has been shown to be lowest in people who had experienced prior infection and vaccination [25]; prior infection alone conferred greater protection than vaccination alone, but both were inferior to hybrid immunity. Given the high level of both prior infection and vaccination coverage in some Brazilian cities, most notably Manaus, it is not surprising that the Delta VOC caused a negligible number of severe cases in these locations (Figures 1D and 3). In Brazil, where vaccination coverage is high and with great spatial heterogeneity in prior attack rates, it makes sense that a marker that reflects immunity from both infection and vaccination (i.e., mean anti-S S/C) is predictive of Delta’s spread.

By November 2021, mean anti-S titres were similarly high across all seven cities. With the introduction of Omicron, there was a large spike in total cases across all cities. However, there was only a modest peak in SARI cases and negligible numbers of deaths. The exception to this was Fortaleza, where a large increase in SARI cases attributable to Omicron infections was observed. However, the decoupling of SARI cases from deaths suggests this was a reporting artifact and not a true reflection of higher morbidity there. Therefore, our results show that antibodies, acquired against historical variants (infection and vaccination), were sufficiently high in Brazil to prevent a significant public health impact of the Omicron VOC. However, due to homogeneity in high mean antibody level across the cities by the time of the Omicron wave, we could not repeat the analysis performed for Delta (Figure 3). If antibody titre data were shared from more varied global locations, the relationship between antibody levels and public health impact (severe cases and deaths) of new variants could be assessed in near real-time and contribute to more informed policy decisions.

### Limitations

There are several limitations to this work. First, we have previously argued that our blood donor samples are representative of the wider population with respect to SARS-CoV-2 infection [6,9,14]. It is less clear to what extent blood donors reflect population vaccination coverage. We can speculate that individuals that donate blood are less likely to be vaccine hesitant than those that do not donate, as blood donation involves engaging with traditional health care services.

Inconsistencies between cities in total reported cases, severe cases (SARI), deaths and attack rate estimation based on blood donor serosurveillence (Figure 1D-E), highlight the ongoing challenges in real-time epidemic monitoring in Brazil. For example, the incidence of total reported cases (Brazilian Ministry of Health) during the Omicron-driven wave varied greatly between cities, with Rio de Janeiro and Fortaleza showing exceptionally large peaks, compared to São Paulo, which increased only slightly. Using alternative metrics of epidemic development may be of value; for example, supplemental figure S4 shows PCR test positivity in a large network of pharmacies located in São Paulo city. Over most of the epidemic, test positivity tracked closely with reported cases, however, in the Omicron-dominated peak, official reported cases appear to have severely underestimated the true magnitude of the epidemic wave. Caution is needed interpreting case data, particularly for this period in Brazil.

Two issues, in particular, are relevant to the application of our results in other settings. Firstly, our study was conducted during vaccine roll out in Brazil, and therefore the findings reflect population antibody titre and immunity before significant waning had occurred. The correlation between protective immunity and antibody titre during the waning phase remains to be assessed at either individual or population levels. If these variables do remain strongly correlated, then monitoring of population antibody titre may be informative about timing of booster doses, for example. Second, it is unclear how mean antibody titres will predict protection against current or future variants that continue to accumulate mutations associated with immune escape and increased fitness [26]. It seems reasonable to assume that mean titres of binding antibodies developed against historical variants would remain correlated with morbidity caused by contemporary or future variants, even if the parameters that describe the relationship are different. For this reason, it is important that serosurveys share antibody titre data, and not simply seroprevalence estimates, so this question can be addressed.

## Data Availability

The serology data required to reporduce the analyses are available at the following repository https://github.com/CADDE-CENTRE. All other data are publically available.

## References

1. Jones JM, Stone M, Sulaeman H, et al. Estimated US Infection- and Vaccine-Induced SARS-CoV-2 Seroprevalence Based on Blood Donations, July 2020-May 2021. JAMA 2021; 326:1400.

2. Bingham J, Cable R, Coleman C, et al. Estimates of prevalence of anti-SARS-CoV-2 antibodies among blood donors in South Africa in March 2022. In Review, 2022. Available at: https://www.researchsquare.com/article/rs-1687679/v2. Accessed 30 May 2022.

3. CDC. Coronavirus Disease 2019 (COVID-19). 2020. Available at: https://www.cdc.gov/coronavirus/2019-ncov/variants/variant-classifications.html. Accessed 25 May 2022.

4. Davies NG, Abbott S, Barnard RC, et al. Estimated transmissibility and impact of SARS-CoV-2 lineage B.1.1.7 in England. Science 2021; 372:eabg3055.

5. Allen H, Vusirikala A, Flannagan J, et al. Household transmission of COVID-19 cases associated with SARS-CoV-2 delta variant (B.1.617.2): national case-control study. The Lancet Regional Health - Europe 2022; 12:100252.

6. Faria NR, Mellan TA, Whittaker C, et al. Genomics and epidemiology of the P.1 SARS-CoV-2 lineage in Manaus, Brazil. Science 2021; 372:815–821.

7. Hoffmann M, Hofmann-Winkler H, Krüger N, et al. SARS-CoV-2 variant B.1.617 is resistant to bamlanivimab and evades antibodies induced by infection and vaccination. Cell Reports 2021; 36:109415.

8. Sabino EC, Buss LF, Carvalho MPS, et al. Resurgence of COVID-19 in Manaus, Brazil, despite high seroprevalence. The Lancet 2021; 397:452–455.

9. Buss LF, Prete CA, Abrahim CMM, et al. Three-quarters attack rate of SARS-CoV-2 in the Brazilian Amazon during a largely unmitigated epidemic. Science 2021; 371:288–292.

10. Crotty S. Hybrid immunity. Science 2021; 372:1392–1393.

11. Khoury DS, Cromer D, Reynaldi A, et al. Neutralizing antibody levels are highly predictive of immune protection from symptomatic SARS-CoV-2 infection. Nat Med 2021; 27:1205–1211.

12. Planas D, Veyer D, Baidaliuk A, et al. Reduced sensitivity of SARS-CoV-2 variant Delta to antibody neutralization. Nature 2021; 596:276–280.

13. Viana R, Moyo S, Amoako DG, et al. Rapid epidemic expansion of the SARS-CoV-2 Omicron variant in southern Africa. Nature 2022; 603:679–686.

14. Junior CP, Buss L, Salomon T, et al. SARS-CoV-2 antibody dynamics in blood donors and COVID-19 epidemiology in eight Brazilian state capitals. In Review, 2022. Available at: https://www.researchsquare.com/article/rs-1363260/v1. Accessed 25 May 2022.

15. Stone M, Grebe E, Sulaeman H, et al. Evaluation of Commercially Available High-Throughput SARS-CoV-2 Serologic Assays for Serosurveillance and Related Applications. Emerg Infect Dis 2022; 28:672–683.

16. OPENDATASUS. Available at: https://opendatasus.saude.gov.br/. Accessed 25 May 2022.

17. LEPP. DEMOGRAFIA | UFRN. 2017; Available at: https://demografiaufrn.net/laboratorios/lepp/. Accessed 25 May 2022.

18. de Souza WM, Buss LF, Candido D da S, et al. Epidemiological and clinical characteristics of the COVID-19 epidemic in Brazil. Nat Hum Behav 2020; 4:856–865.

19. Brizzi A, Whittaker C, Servo LMS, et al. Spatial and temporal fluctuations in COVID-19 fatality rates in Brazilian hospitals. Nat Med 2022; Available at: https://www.nature.com/articles/s41591-022-01807-1. Accessed 27 May 2022.

20. GISAID - Initiative. Available at: https://www.gisaid.org/. Accessed 25 May 2022.

21. Venables W, Ripley B. Modern Applied Statistics with S, Fourth edition. Springer, New York. Available at: https://www.stats.ox.ac.uk/pub/MASS4/.

22. Earle KA, Ambrosino DM, Fiore-Gartland A, et al. Evidence for antibody as a protective correlate for COVID-19 vaccines. Vaccine 2021; 39:4423–4428.

23. MendronelJunior A, Dinardo CL, Ferreira SC, et al. Correlation between SARSlCOVl2 antibody screening by immunoassay and neutralizing antibody testing. Transfusion 2021; 61:1181–1190.

24. Otter AD, D’Arcangelo S, Whitaker H, et al. Determinants of SARS-CoV-2 anti-spike antibody levels following BNT162b2 vaccination: cross-sectional analysis of 6,000 SIREN study participants. Infectious Diseases (except HIV/AIDS), 2022. Available at: http://medrxiv.org/lookup/doi/10.1101/2022.04.21.22274025. Accessed 25 May 2022.

25. Goldberg Y, Mandel M, Bar-On YM, et al. Protection and Waning of Natural and Hybrid Immunity to SARS-CoV-2. N Engl J Med 2022; :NEJMoa2118946.

26. Obermeyer F, Jankowiak M, Barkas N, et al. Analysis of 6.4 million SARS-CoV-2 genomes identifies mutations associated with fitness. Science 2022; :abm1208.

