## Supplemental Material for "Predicting SARS-CoV-2 variant spread in a completely seropositive population using semi-quantitative antibody measurements in blood donors"

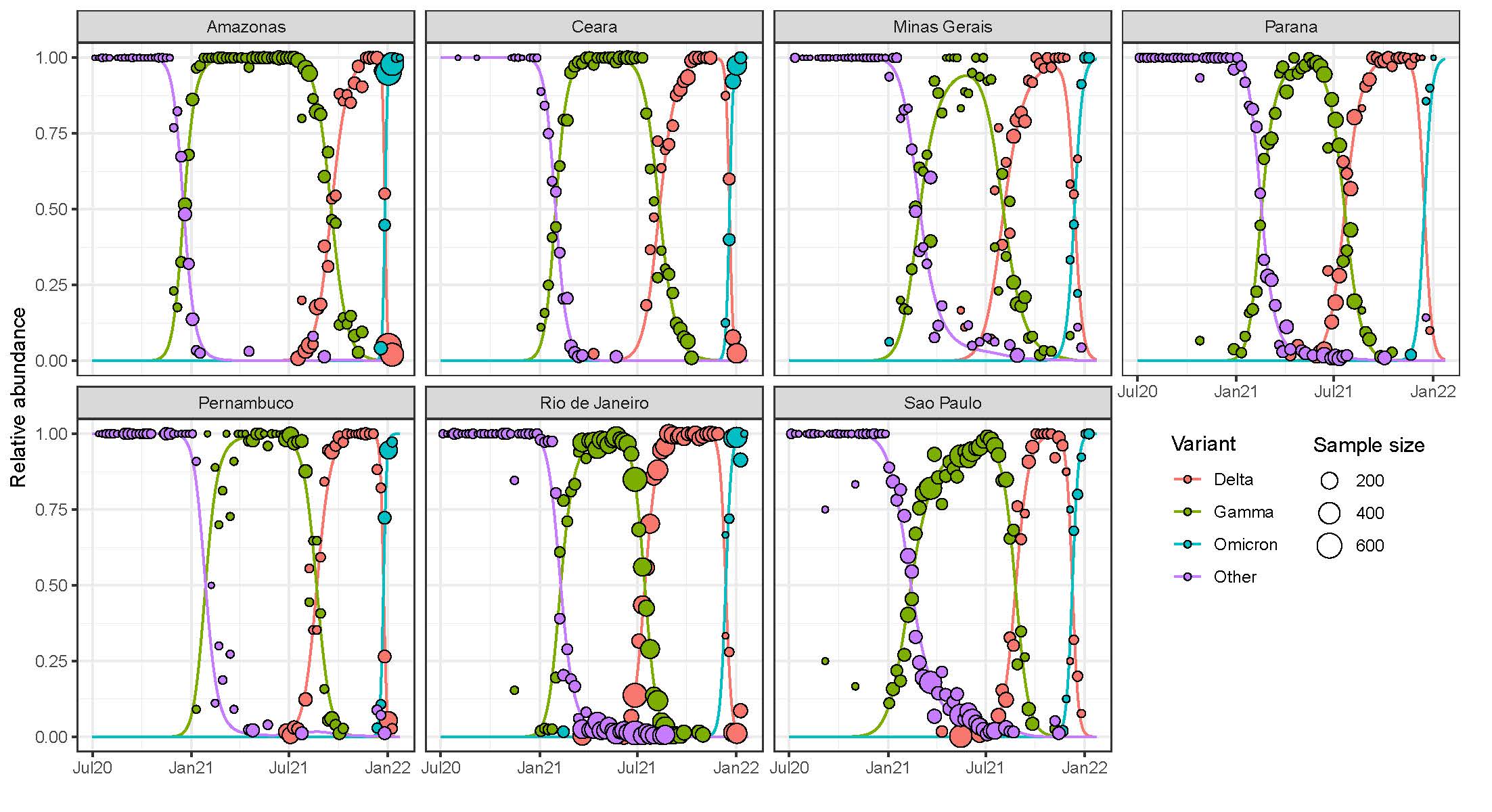

**Supplemental Figure S1.** The metadata for all SARS-CoV-2 sequences deposited on GISAID between Jul 2020 and Jan 2022 (<https://www.gisaid.org/>) were downloaded. The lines show the predictions of a multinomial model fit using the nnet package in R. “Other” refers principal to wild type virus and P.2.

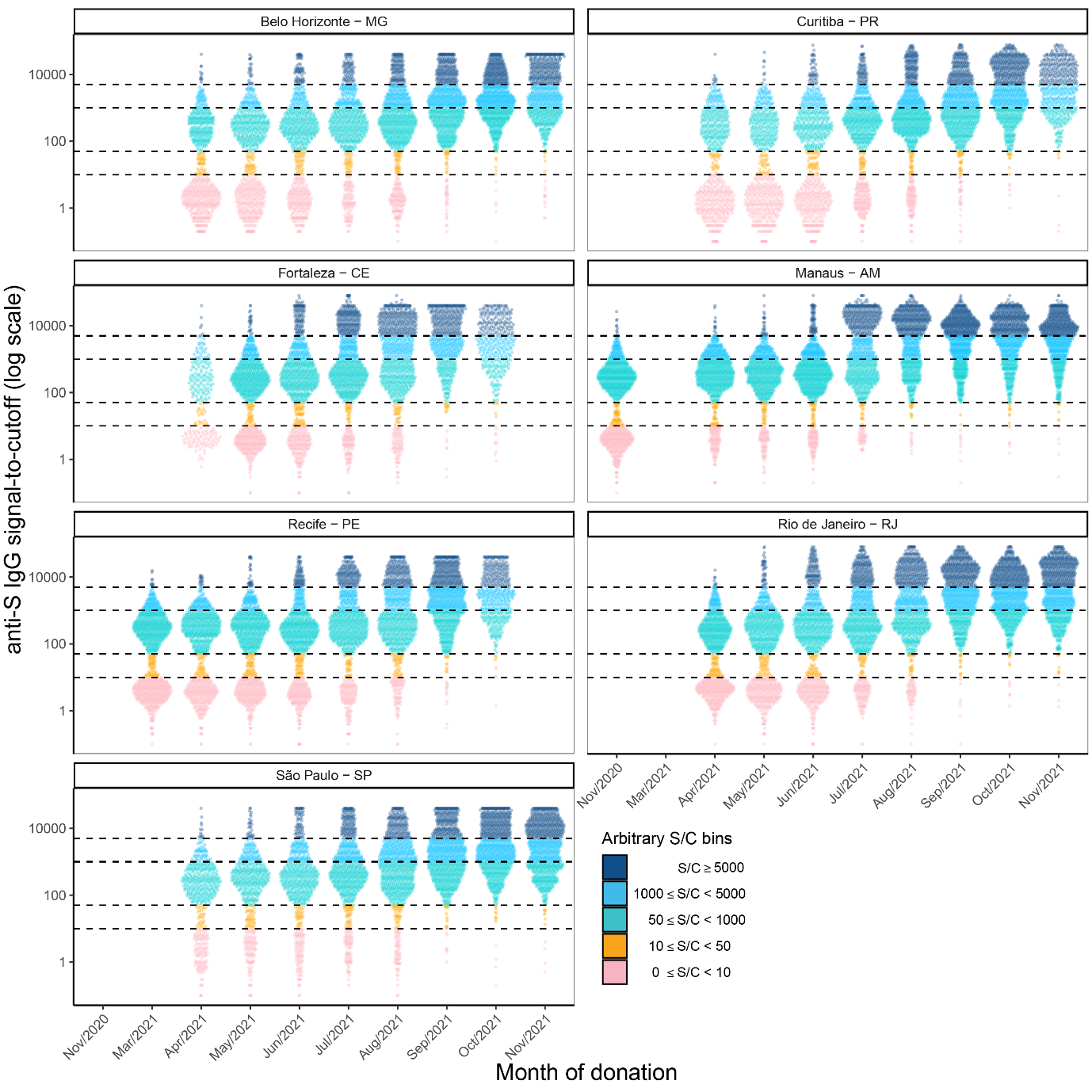

**Supplemental Figure S2.** Raw signal to cut-off readings for each month blood donor sample across the seven Brazilian state capital cities. A threshold of 50 S/C is defined as a positive assay, other thresholds are arbitrary and shown to aid visualization.

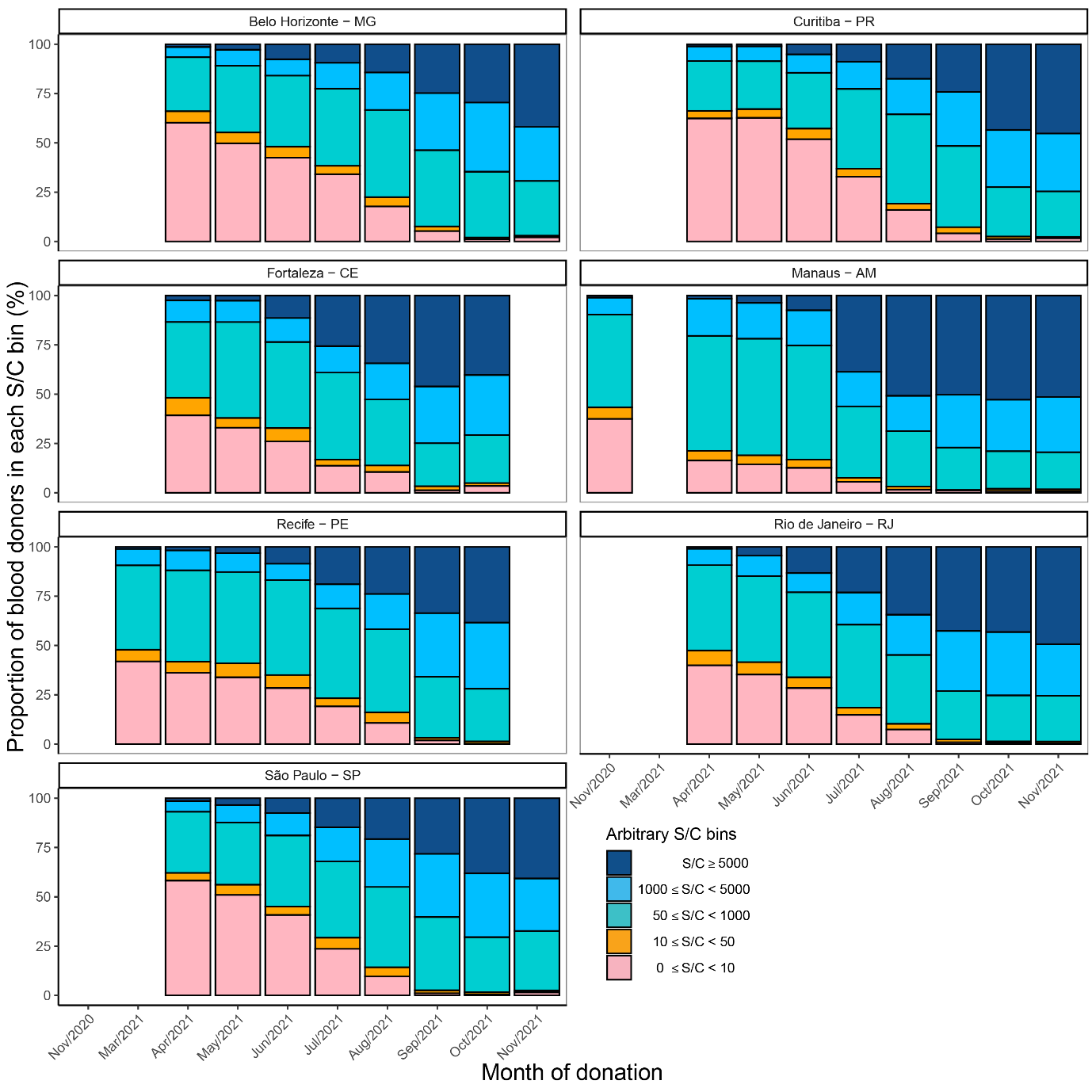

**Supplemental Figure S3.** Proportion of blood donor samples falling in arbitrary S/C bins each month across seven Brazilian state capitals.

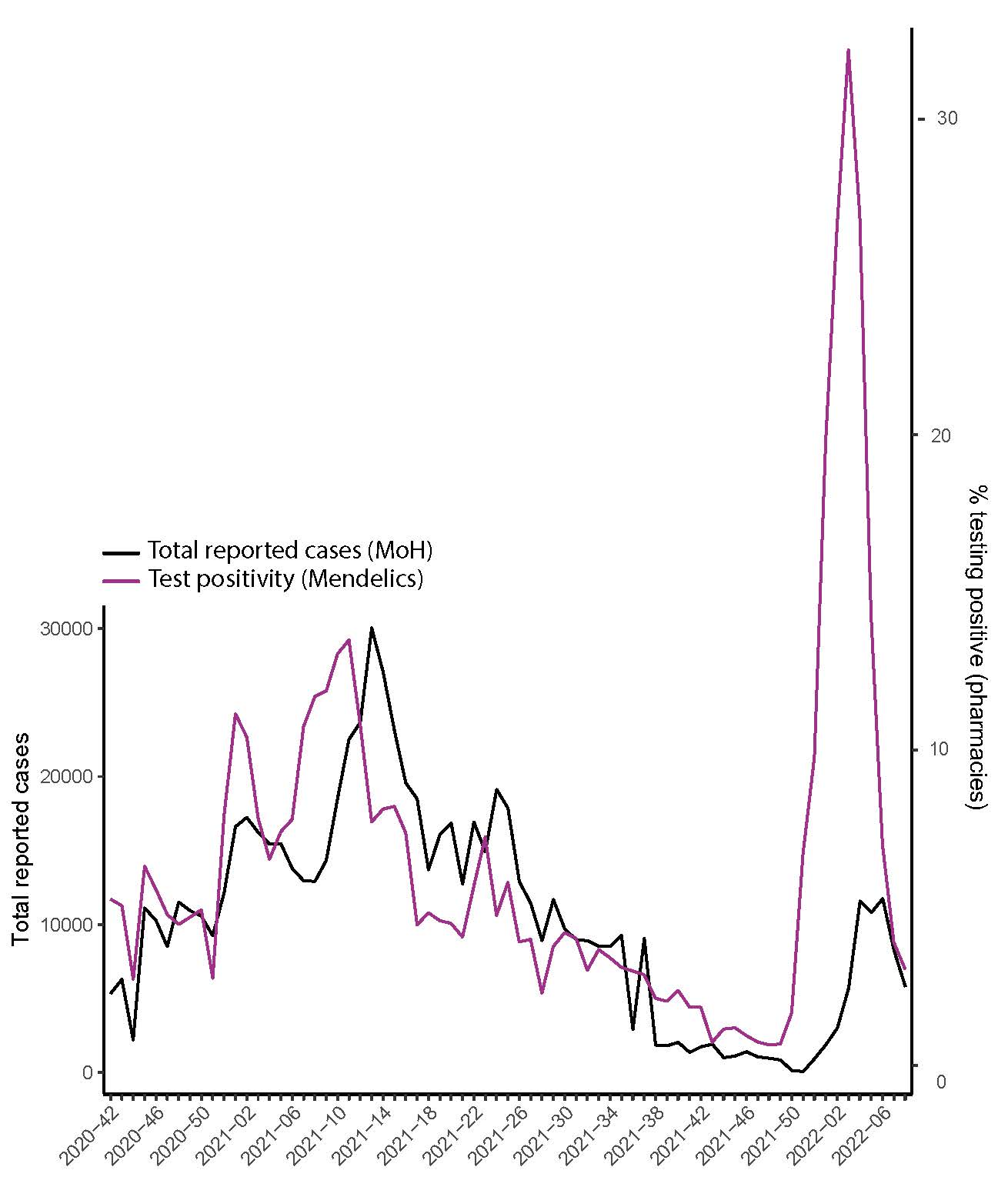

**Supplemental Figure S4.** Black line (LHS y-axis) **-** total number of reported cases in Sao Paulo city (https://[www.covid.saude.br](http://www.covid.saude.br)/) shown per week of reporting. Purple line (RHS y-axis) shows the % positivity of tests administered in pharmacies in Sao Paulo by Mendelics (https://mendelics.com.br/).
